# Linking Data-Driven Symptom Dimensions to Resting-State EEG Biomarker Candidates in Chronic Pain

**DOI:** 10.64898/2026.01.23.26344711

**Authors:** Paul Theo Zebhauser, Felix S. Bott, Enayatullah Baki, Henrik Heitmann, Nicolo Bruna, Elisabeth S. May, Markus Ploner

## Abstract

Chronic pain (CP) is a multidimensional condition characterized by physical, emotional, and cognitive symptoms. However, many neuroimaging studies investigating the brain mechanisms of CP have focused on single-domain measures, most commonly pain intensity. Incorporating multidimensional symptom profiles may advance the understanding of CP, its neural underpinnings, and the development of clinically actionable biomarkers. Here, we aimed to empirically derive symptom dimensions of CP and relate them to resting-state brain activity and connectivity measured by electroencephalography (EEG). Using a data-driven approach, we identified latent symptom dimensions in 207 individuals with CP based on the brief, internationally validated PROMIS-29 profile, which assesses general health across key physical, mental, and social domains. Principal component analysis revealed two dimensions, affective burden and physical burden, closely corresponding to established PROMIS-derived health dimensions in other clinical populations. Resting-state EEG was obtained in a subsample of 116 participants using a mobile, rapid-to-deploy 29-channel dry-electrode system. Bayesian regression analyses provided moderate to strong evidence for a negative association between affective burden and beta-band connectivity, particularly in left frontal and somatomotor regions. Together, these findings demonstrate how empirically derived symptom dimensions captured by PROMIS-29 can be linked to scalable, network-level EEG biomarkers. This framework illustrates an EEG-informed strategy for biopsychosocial stratification in CP, with potential relevance for personalized symptom-targeted interventions.

## 1. Introduction

Chronic pain (CP) is best understood within a biopsychosocial framework [10]. Accordingly, people with CP typically experience complex patterns of physical, affective, and cognitive symptoms alongside persistent pain, resulting in substantial impairments in quality of life [6]. Understanding these symptom patterns is crucial for gaining mechanistic insights and for developing personalized treatment strategies. Brain function plays a central role in CP [21], and identifying brain-based biomarkers that reflect symptom burden could improve assessment, stratification, and individualized treatment. Consequently, many neuroimaging studies have investigated the relationship between selected symptoms, most often pain intensity, and brain function. Broadening the scope of such studies from single symptoms to symptom patterns may advance the understanding of the brain mechanisms of CP and naturally motivates the development of *dimensional* brain biomarkers that reflect underlying symptom dimensions rather than individual symptoms.

Recent advances in neuroscience increasingly favor a view of brain disorders characterized by continuous variation in symptom patterns and biological features, both within specific disorders and across traditional diagnostic boundaries [18]. Rather than mapping onto discrete symptoms or diagnostic categories, alterations in brain function may therefore reflect variation along latent symptom dimensions. Several lines of reasoning support this view. First, the functional organization of the brain is inherently network-based [29], with large-scale networks linked to broad functional domains rather than specialized subfunctions [25; 42]. Correspondingly, disturbances of these networks tend to produce clinical manifestations that align with broad symptom dimensions rather than single symptoms [18]. Second, broad symptom dimensions are more stable over time than state-like symptoms. For example, pain intensity fluctuates considerably in CP [33], whereas broader constructs, such as affective distress, remain relatively stable over time [12; 16] and thus are more likely to map onto resting-state brain activity. Third, dimensional brain biomarkers can meaningfully support personalized interventions, including neuromodulation, cognitive-behavioral strategies, and pharmacological treatments, by enabling stratification into clinically relevant subgroups [9; 38].

Electroencephalography (EEG) is a particularly promising tool in this context. With recent advances in hardware and analysis methods, EEG offers a direct and scalable measure of brain function. Numerous studies have demonstrated EEG correlates of pain in people with CP, representing initial steps towards clinically useful biomarkers [3; 40; 44].

Here, we aimed to advance these efforts by adopting a dimensional approach to CP and relating broad symptom dimensions to brain function. We sought to (i) empirically identify symptom dimensions of CP using a brief, widely available, and validated patient-reported outcome (PRO) tool and (ii) relate these dimensions to resting-state brain function. To this end, we administered the PROMIS-29 [14] to a large sample of people with CP and used Principal Component Analysis (PCA) to derive latent symptom dimensions (Fig. 1). We then examined their associations with resting-state EEG measures of brain function (Fig. 1). This study thus extends EEG biomarker research in CP by moving from unidimensional indicators (e.g., pain intensity) to multidomain symptom dimensions, supporting biopsychosocial stratification, outcome prediction, and the development of targeted interventions.

**Figure 1.**
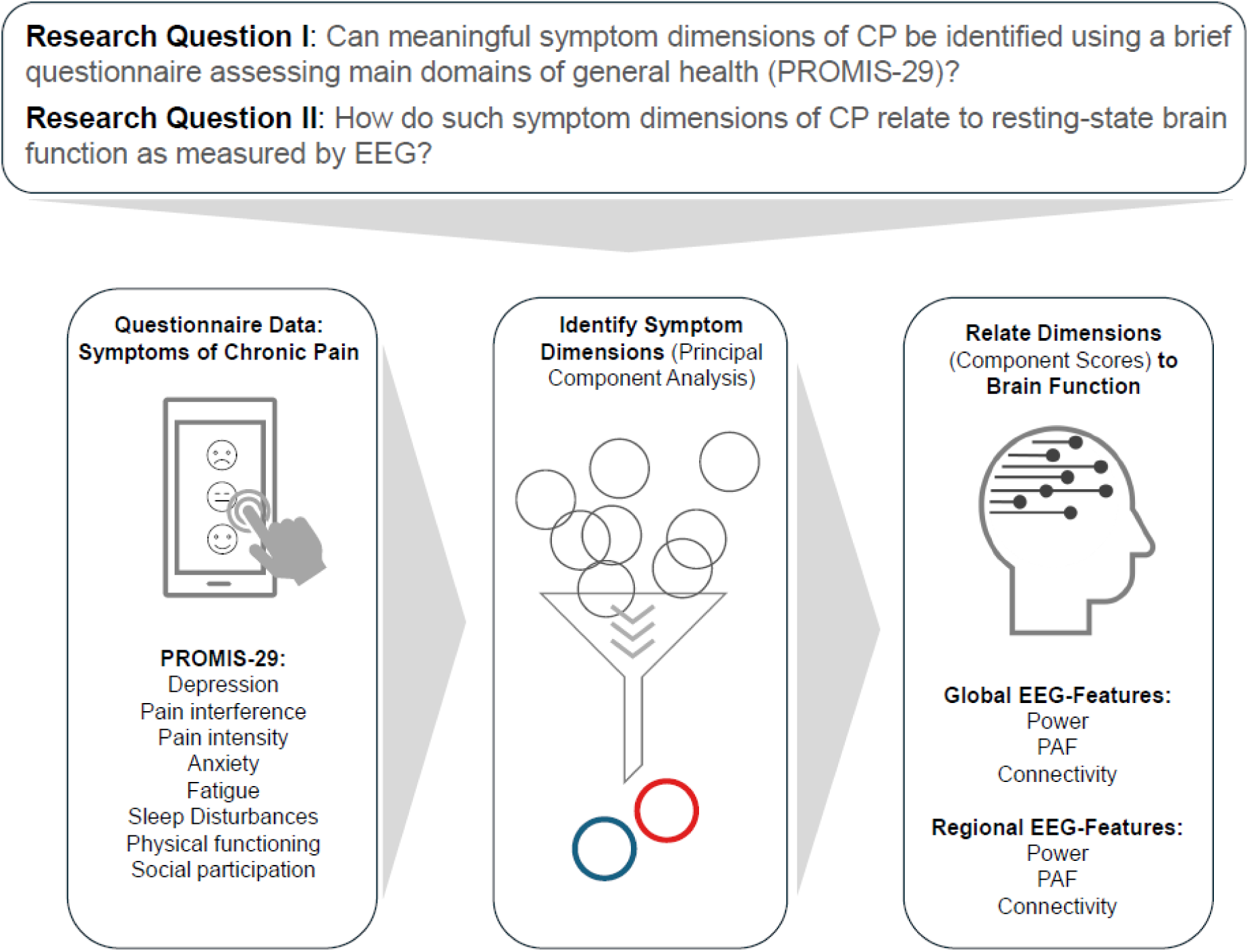
Overview of study design. CP = chronic pain, PAF = peak alpha frequency.

## 2. Methods

The study protocol and analysis plan were preregistered (https://osf.io/7bwnh/registrations), and data and code for analyses will be made publicly available upon publication of this manuscript (https://osf.io/7bwnh/files). The study protocol was approved by the Ethics Committee of the Medical Faculty of the TUM and conducted following the latest version of the Declaration of Helsinki.

### 2.1. Study Participants

All study participants were adults (minimum age of 18 years) with CP, defined as pain lasting a minimum of three months. They were recruited through convenience sampling from the outpatient clinic of the *Center for Interdisciplinary Pain Medicine* at the *TUM University Hospital Rechts der Isar,* Munich, Germany. For the present project, these data were integrated with questionnaire data from an ongoing study on anhedonia in CP (https://osf.io/fpnqw), conducted at the same site. In total, 207 participants with CP completed questionnaire data on CP symptoms, of whom 120 participants underwent resting-state EEG recordings. Individuals were excluded from the EEG study if they had severe concomitant neurological (e.g., multiple sclerosis) or psychiatric (e.g., post-traumatic stress disorder, schizophrenia) disorders, a primary headache disorder, or reported regular or recent intake of benzodiazepines. Other centrally acting medications were not considered an exclusion criterion, as a recent study provided evidence against effects on EEG metrics [43]. Three participants with EEG recordings were excluded from all analyses due to missing questionnaire data resulting from a technical error during data transfer from the recording tablet to storage. One participant was excluded from analyses after it was determined that they had suffered a stroke prior to study participation.

### 2.2. Clinical and Questionnaire Data

We administered the PROMIS-29v2.1 [14] to a sample of 207 people with CP of mixed etiologies. The PROMIS-29 is a multilingually available, validated patient-reported outcome measure designed to capture core determinants of physical, mental, and social health, which are central contributors to quality of life across medical conditions. By assessing these broad health domains in a standardized and transdiagnostic manner, the PROMIS-29 is particularly well-suited for multifaceted conditions, such as CP. Its multidimensional structure assesses depression, anxiety, fatigue, pain interference, pain intensity, physical functioning, sleep disturbance, and social participation across 29 items rated on 5-point Likert scales. For all symptom domains, sum scores (4 items per domain, ranging from 4 to 20, excluding pain intensity, which has only one item) were calculated. Furthermore, T-values were derived based on T-score conversion tables (using the PROMIS-29 Profile v2.1 scoring manual).

### 2.3. Principal Component Analysis of Questionnaire Data

To derive latent symptom dimensions from questionnaire data assessed by the PROMIS-29, PCA using Varimax rotation was conducted using the 8 symptom domain scores (sum scores for depression, anxiety, fatigue, pain interference, physical functioning, sleep disturbance, social participation, and pain intensity). This approach was adapted from a recent PCA-based analysis of psychopathological symptoms and personality dimensions of people with CP [39]. Scores of physical functioning and social participation were inverted beforehand, as higher scores indicate better functioning for those scales. Analyses were performed in *R* v4.4 [36] using the *psych* package. Components were retained using the Kaiser criterion (eigenvalues>1, thereby keeping those components that explain more variance than a single variable), confirmed by inspection of the scree plot using the elbow method. Component loadings of questionnaire scales were examined to ensure the interpretability of latent dimensions. Individual component scores were extracted for subsequent analyses (e.g., EEG–symptom associations) by projecting each participant’s standardized variable values onto the retained principal components. This resulted in a set of continuous scores for each individual, reflecting the extent to which each principal component was represented in that individual’s symptom profile. These scores were then used in subsequent analyses to examine relationships with other variables (demographics and EEG features).

### 2.4. Associations of Component Scores with Demographics

Associations of component scores derived using PCA with gender were examined using Bayesian t-tests and the *BayesFactor* package in R (default JZS prior: Cauchy 0, 0.707); effect sizes were estimated using Cohen’s d. Correlations of component scores with age were computed using the *BayesFactor* package in R (default Jeffrey’s adjusted beta prior); effect sizes were estimated using Pearson’s correlation coefficients. Bayes Factors (BF10) were interpreted as strong, moderate, or weak evidence (>10, 3–10, >1<3 regarding the alternative hypothesis; < 0.1, >0.1<0.33, 0.33–1 regarding the null hypothesis; [41]).

### 2.5. EEG Data Acquisition and Preprocessing

Participants were seated comfortably in a quiet room and instructed to remain relaxed but awake. Resting-state EEG was recorded in 5-minute eyes-open and eyes-closed conditions; only eyes-closed recordings were used for subsequent analyses. EEG was acquired using a dry electrode system with 29 channels (CGX Quick 32r, San Diego, CA) with a wireless amplifier. The electrode layout included all standard 10–20 system electrodes, as well as additional electrodes Fpz, AF7/8, FC5/6, CP5/6, PO7/8, and Oz. During recording, signals were referenced and grounded at A1 (left earlobe). Data was sampled at 500 Hz, and electrode impedances were maintained below 2500 kΩ, consistent with the higher impedance levels typically observed in dry EEG systems compared to conventional wet systems [20].

EEG data were preprocessed and analyzed in MATLAB using the openly available DISCOVER-EEG pipeline [15] with the EEGLab [11] and FieldTrip [27] toolboxes. Preprocessing (default settings) included line noise removal, bad channel rejection, re-referencing, independent component analysis, bad component removal, bad channel interpolation, and bad segment removal.

### 2.6. EEG Analysis

We analyzed a comprehensive set of EEG activity and connectivity features typically used in CP research, both across the entire brain (global features) and in specific regions (regional features). To assess the distributional properties of EEG features, we applied the Kolmogorov–Smirnov test. Variables with a KS p-value below 0.05 were considered significantly deviating from a normal distribution and were tested for direction of skewness. As all non-normally distributed variables (mainly power metrics) were right-skewed and non-negative, a log transformation was applied. Global power and peak alpha frequency (PAF) were calculated in sensor space; all connectivity and region-specific metrics were calculated in source space.

Source reconstruction of the preprocessed EEG data was performed using an array-gain linear constrained minimum variance (LCMV) beamformer [11]. Data were then projected to source space defined by the 100-parcel Schaefer atlas (7-network version) [32]. Lead fields were computed using a volume conduction model based on the Montreal Neurological Institute (MNI) template (standard_bem.mat) in FieldTrip. Spatial filters were derived from covariance matrices of band-pass filtered data and the lead fields, with 5% regularization applied to mitigate rank deficiencies. Functional connectivity was estimated between all reconstructed virtual time series for each frequency band

For all analyses, frequency bands were defined according to the *COBIDAS MEEG* recommendations [28]: theta (4 to < 8 Hz), alpha (8 to < 13 Hz), beta (13 to 30 Hz), and gamma (> 30 to 80 Hz).

Global PAF was calculated as the center of gravity (CoG; of the alpha spectrum (weighted average of alpha band frequencies, with each frequency weighted by its power), averaged across all electrodes (channels in sensor space. Global power was calculated by averaging absolute power values for defined frequency bands across channels in sensor space. Global connectivity per frequency band was quantified as the amplitude envelope correlation coefficients (AEC) averaged across all pairwise source-level connections (upper triangle of connectivity matrix), a standard approach in EEG research [4; 5; 15; 37]. All regional EEG features were calculated in the above-specified source space, using the 100-parcel Schaefer atlas [32]. Cortical region labels for interpretation were assigned using the *BioImage-Suite-webtool* (https://bioimagesuiteweb.github.io/webapp/mni2tal.html) with Talairach Atlas labels [22; 23], based on MNI coordinates from the Schaefer atlas. Spatially resolved analogues of global EEG features were considered for region-specific analyses. For regional power, the source absolute power per parcel, as provided by DISCOVER-EEG [11], was used. For regional PAF, the CoG of the alpha spectrum was calculated in source space. For regional connectivity, mean connectivity (AEC) for single parcels with all other parcels was calculated, often referred to as node strength [31].

### 2.7. Regression Analysis of Symptom Dimensions and EEG Features

fWe used a default JZS prior on the regression coefficients (r-scale = 0.354) and a binomial model prior with α = β = 1. Effect sizes for individual predictors were estimated as the squared standardized regression coefficients (*r2*) reflecting the proportion of variance each predictor uniquely explained. As Bayesian inference provides probabilistic estimates for each effect, frequentist-style corrections for multiple tests were not applied [2; 34]. Therefore, no techniques for correcting multiple testing were applied. A power analysis for a multiple regression model with four predictors (two component scores, age, and gender), assuming a significance level (α) of 0.05 and statistical power (1 – β) of 0.80, indicated that our sample size (n=116) was sufficient to detect a small-to-medium effect (Cohen’s f² = 0.11).

## 3. Results

### 3.1. Clinical Characteristics

Table 1 presents the demographic and clinical characteristics, as well as the domain sum scores obtained using the PROMIS-29, of both the full sample and the EEG subsample. Fig.2 illustrates the percentage of clinically relevant symptoms (indicated by T-values greater than 1) in PROMIS domains across the full sample. Those ranged from 19% for sleep interference to 69% for pain interference. Psychopathological symptoms of depression, anxiety, and fatigue were found to be in a clinically meaningful range for 40, 35, and 42%, respectively. Fig. 3 shows the distribution of CP diagnoses across both samples.

**Figure 2.**
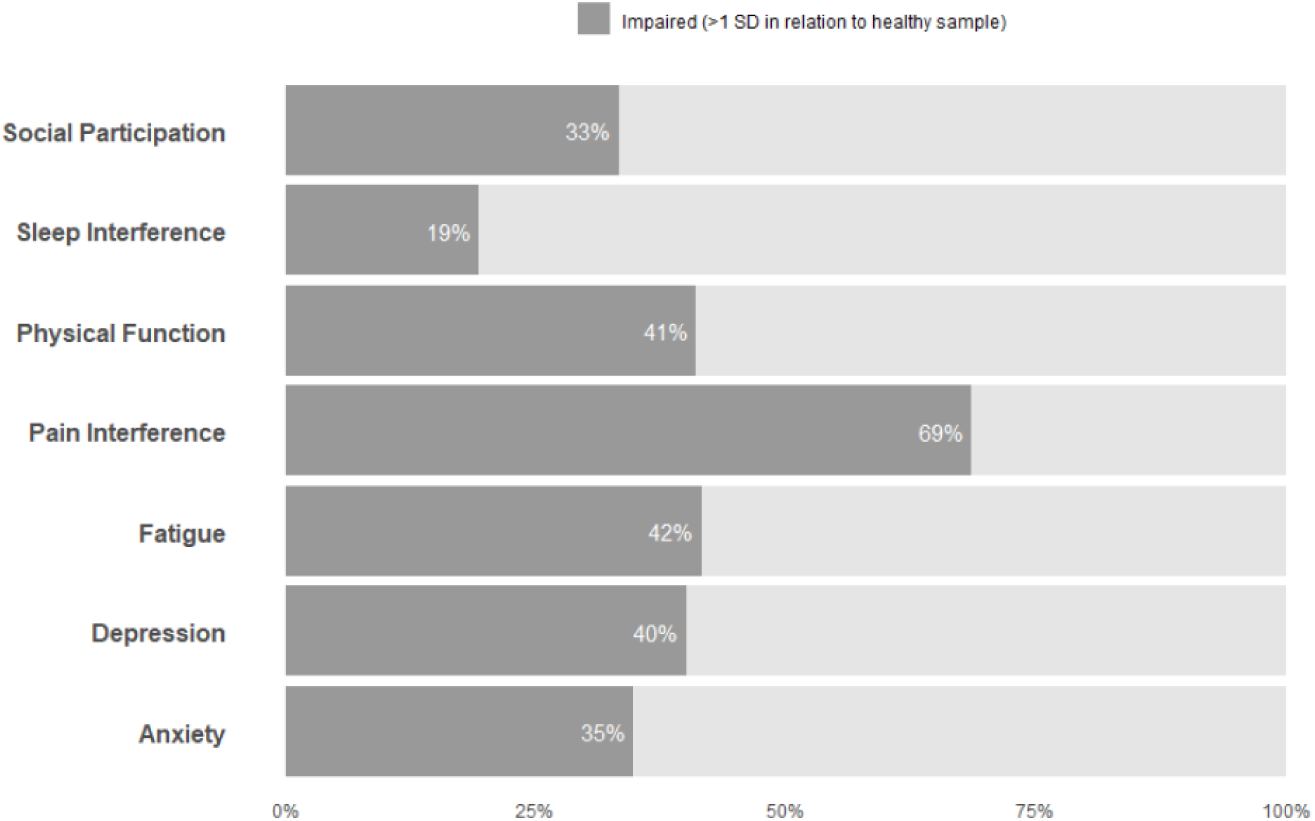
Prevalence of clinically relevant symptoms (full sample). SD = Standard Deviation

**Fig. 3.**
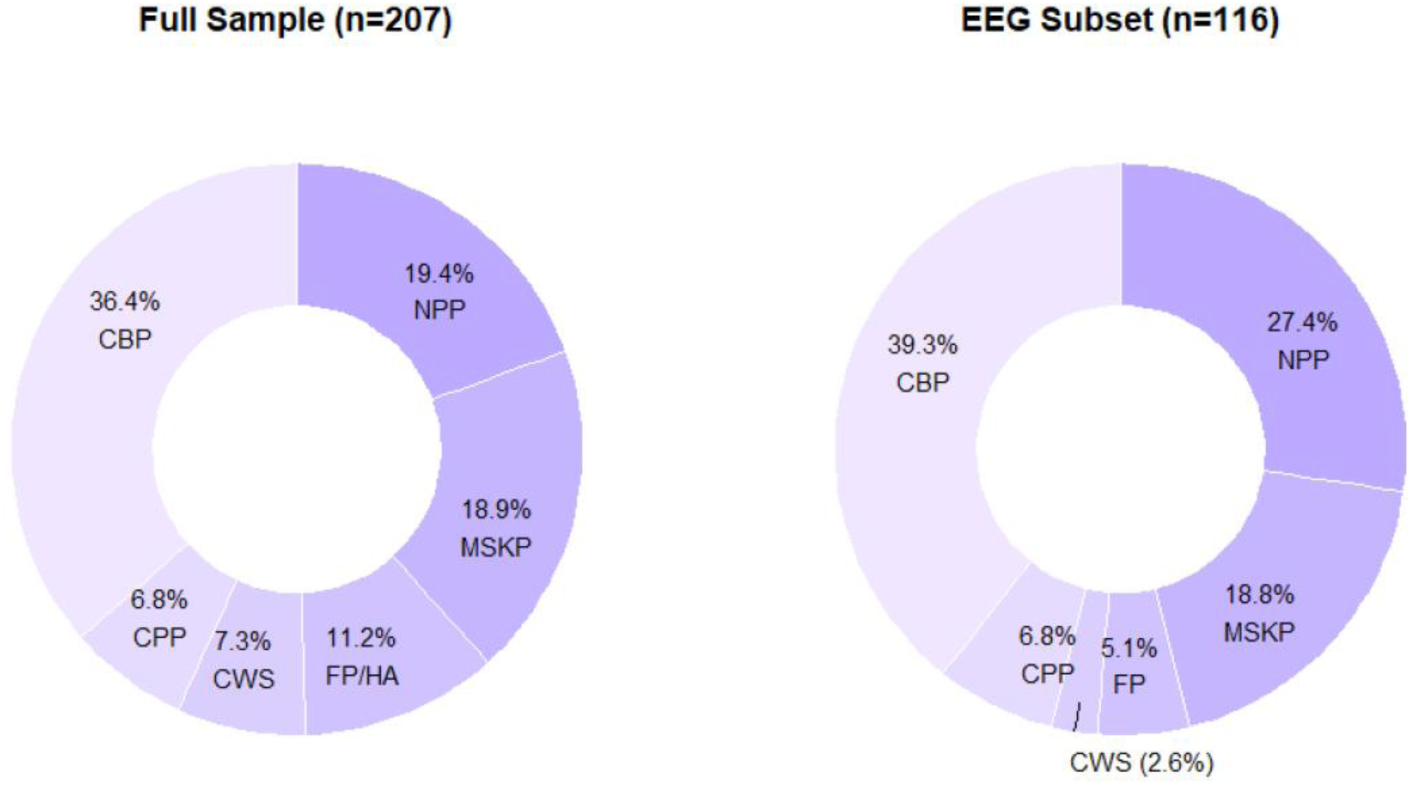
Distribution of chronic pain diagnoses in the full and the EEG subsample. Note that in the EEG subsample, people with headache conditions were excluded. CBP = Chronic Back Pain, NPP = Neuropathic Pain, MSKP = Other Musculoskeletal Pain, FP/HA = Facial Pain/Headache, CWS = Chronic Widespread Pain, CPP = Chronic Pelvic Pain.

**Table 1.** Demographic and Clinical Characteristics of Study Participants.

|  | Full Sample | EEG-Subsample |
| --- | --- | --- |
| **n** | 207 | 116 |
| **Female (%)** | 63.6% | 56% |
|  | *Mean (SD)* | *Mean (SD)* |
| **Age (years)** | 53.4 (17.1) | 56.8 (17.1) |
| **Pain Intensity (0-10)** | 5.6 (2.1) | 5.3 (2.3) |
| **Pain Duration (months)** | 102.4 (116.0) | 86.2 (100.8) |
| **Physical Function\* (4-20)** | 15.0 (4.0) | 15.0 (4.3) |
| **Anxiety (4-20)** | 9.3 (3.6) | 8.2 (3.1) |
| **Depression (4-20)** | 9.6 (3.5) | 8.6 (3.3) |
| **Fatigue (4-20)** | 12.4 (4.2) | 11.2 (4.1) |
| **Sleep Interference (4-20)** | 11.9 (4.0) | 11.4 (4.0) |
| **Social Participation\* (4-20)** | 11.6 (4.2) | 12.4 (4.5) |
| **Pain Interference (4-20)** | 13.0 (4.0) | 12.6 (4.2) |

\*Higher values indicate better functioning, SD = Standard Deviation

### 3.2. Principal Component Analysis

Two components (see Fig. 4) were retained based on the criteria defined above. Component 1 explained 35.1% of the variance. As it was characterized primarily by high loadings on anxiety (0.886), depression (0.844), and fatigue (0.720), it was labeled “affective burden”. Component 2 explained 31.9% of the variance and was characterized mainly by loadings on pain interference (0.831), physical function (0.850), and pain intensity (0.692). It was labeled “physical burden”. Together, the two components accounted for 66.9% of the total variance, which is considered strong for PCA of questionnaire data [13].

**Fig. 4.**
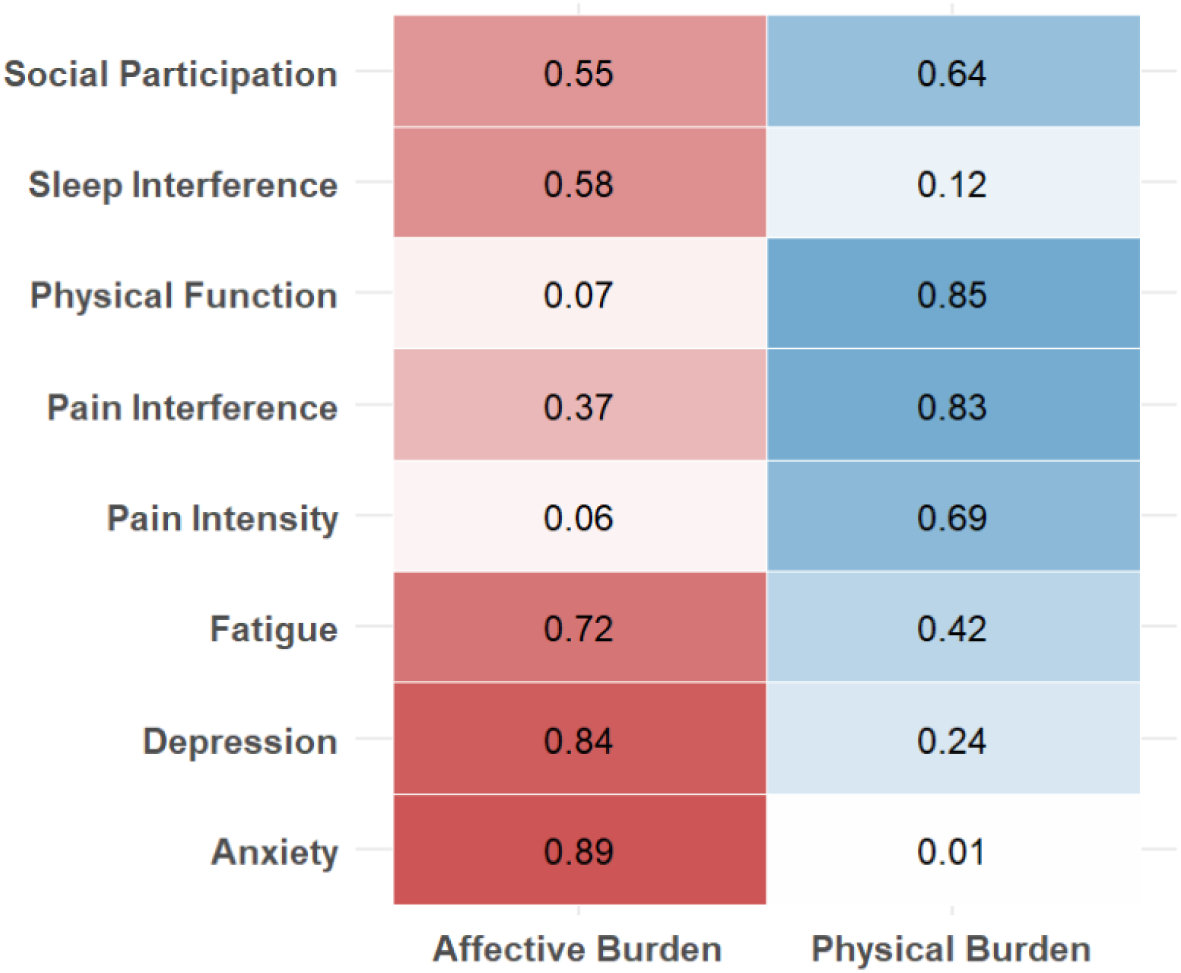
Results of the Principal Component Analysis. Values in cells and color intensity indicate variable loadings on retained components (higher color intensity = higher loading). The two components accounted for 66.9% of the total variance (35.1% for affective, 31.9% for physical burden).

### 3.3. Symptom Dimensions and Demographics

We examined associations between component scores and age and gender. We found strong evidence for a negative correlation of affective burden with age (r = - 0.33, BF10 > 103) and moderate evidence for a positive correlation of physical burden with age (r = 0.19, BF10 = 5.3), indicating lower affective but higher physical burden with increasing age (see Fig. 5). Furthermore, Bayesian t-Tests provided strong evidence for higher scores of affective burden for female participants (BF10 = 50.4, d = 0.51, see Fig. 6). In contrast, we found weak evidence against a gender-difference in physical burden (BF10 = 0.7, d = 0.26).

**Fig. 5.**
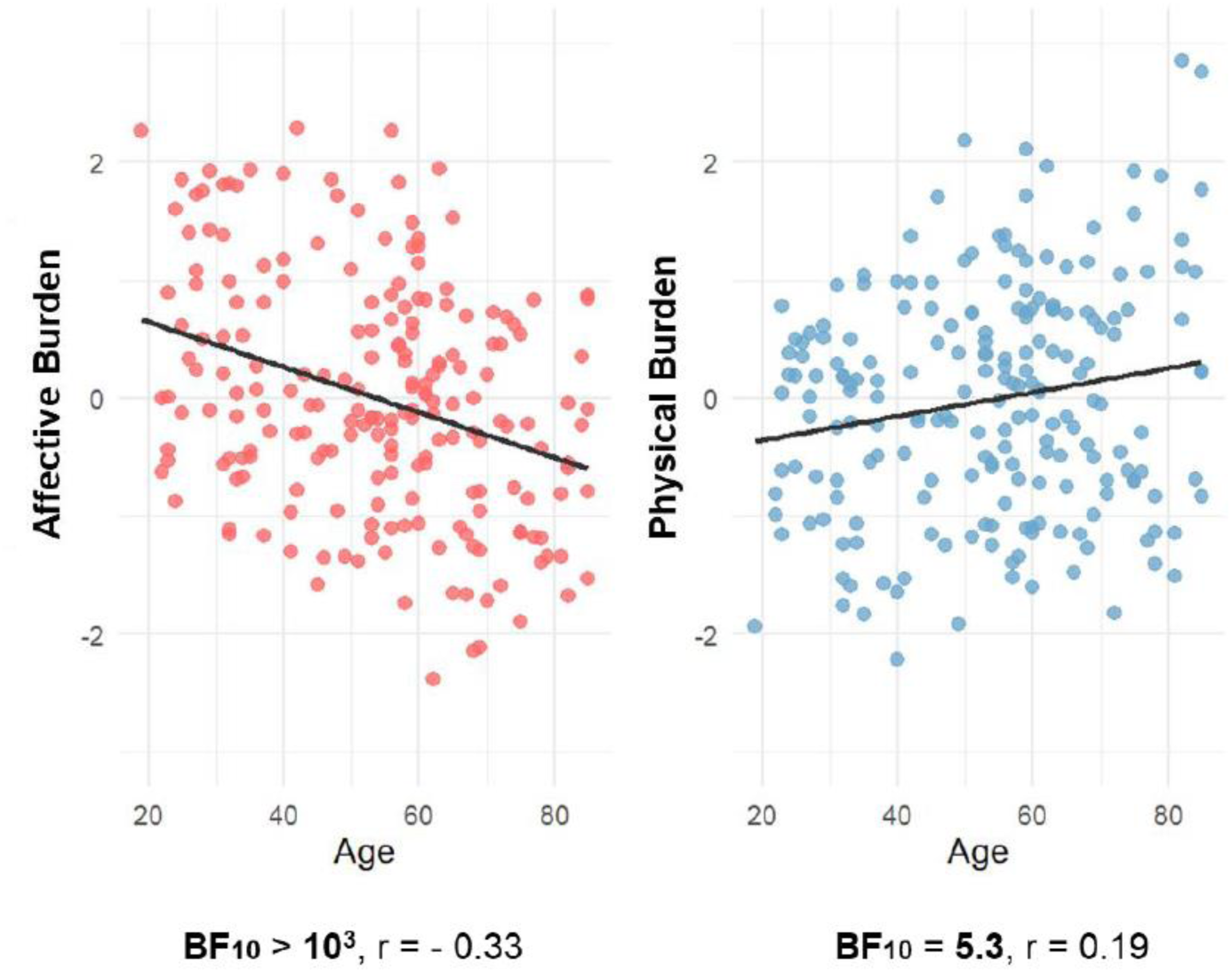
Scatter plots showing associations of scores for affective and physical burden with age. Note that age was negatively correlated with affective burden, while a (weaker) positive association was observed for age and physical burden. BF = Bayes factor. Solid lines indicate fitted regression lines.

**Fig. 6.**
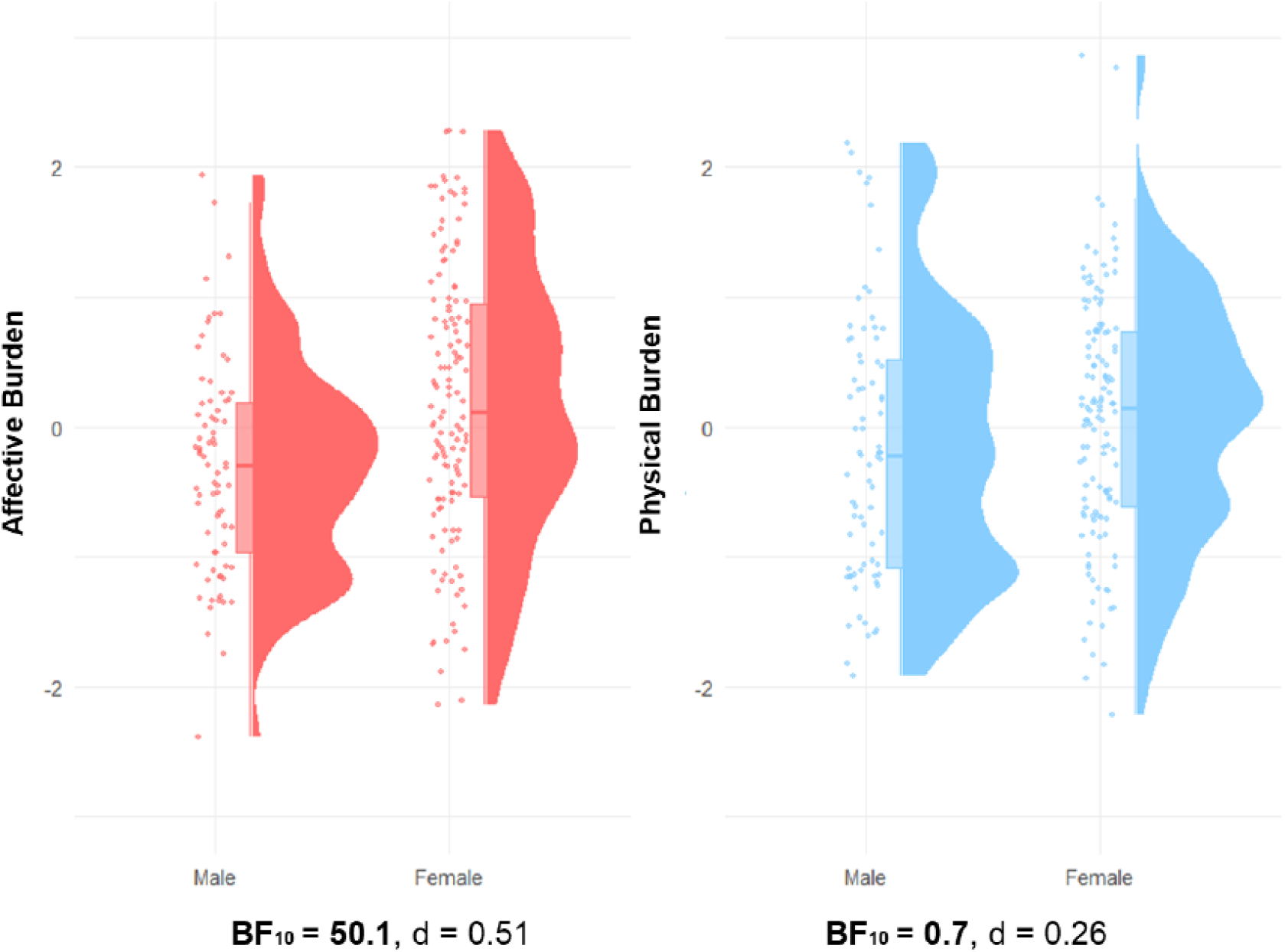
Raincloud plots of scores for affective and physical burden, split by gender. Raincloud plots display individual data points, boxplots, and probability density functions. The box in the boxplot extends from the first to the third quartile, with a thicker line indicating the sample median. Note that female participants scored higher on the affective burden component, while no differences were observed for physical burden. BF = Bayes factor, d = Cohen’s d.

### 3.4. Regression Analysis of Symptom Dimensions and EEG Features

Overall, robust EEG associations were confined primarily to beta-band connectivity, whereas associations with power and other frequency bands were weak or inconsistent. Full results for all regression models, BFs for inclusion of predictors, posterior means with standard deviations, effect size estimates, and frequentist p-values are provided in Supplement S1. In the following sections, we present a summary and selected results.

For global EEG features (see Fig. 7), we found moderate evidence for a negative association between affective burden and beta connectivity (BF10 = 4.91, r^²^ = 0.03 *[small to medium effect]*), indicating an association of higher affective burden with lower beta connectivity. Gamma power and beta connectivity were very weakly associated with physical burden. For all remaining global EEG features, we found weak to moderate evidence against associations with physical and affective burden.

**Fig. 7.**
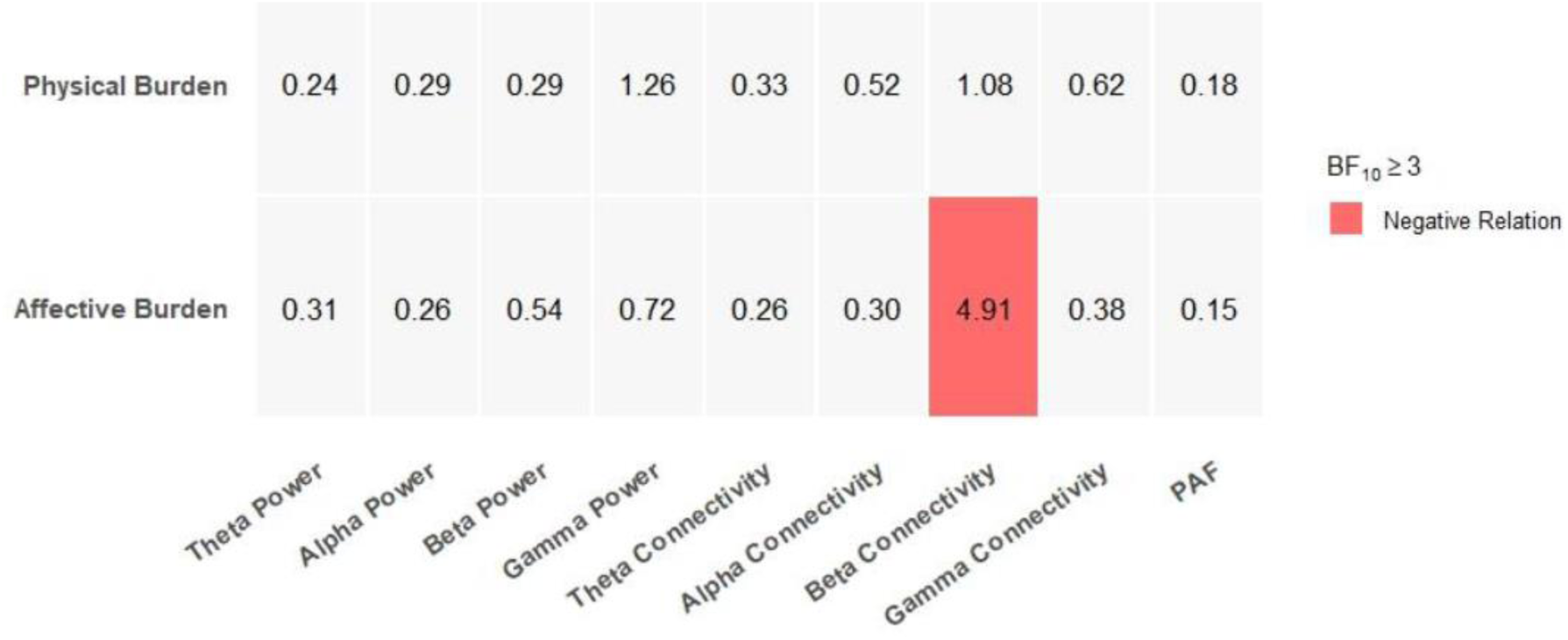
Overview of regression analyses of affective/physical burden on global EEG features. Only BFs indicating at least moderate evidence for effects are plotted. PAF = peak alpha frequency. BF = Bayes Factor.

Regarding regional/local EEG features, effects were also mostly confined to beta-band connectivity. In line with the global findings, we found moderate (18 parcels mainly distributed in left somatomotor and frontal regions; BF10 = 3.0-9.2, r^2^ = 0.01-0.04 *[small to medium effects]*) to strong (4 parcels across left somatomotor regions; BF10 = 13.0-40.1, r2 = 0.05-0.08 *[small to medium effects]*) evidence for negative associations of regional beta connectivity with affective burden (see Fig. 8). In adjacent regions, we found weak evidence for the same association. In addition, we found moderate (4 parcels mainly distributed in right frontal regions; BF10 = 3.5-9.8, r^2^ = 0.02-0.04 *[small to medium effects]*) to strong (1 parcel in the left prefrontal cortex; BF10 = 31.7, r^2^ = 0.06 *[small to medium effect]*) evidence for negative associations with physical burden (see Fig. 8), again with weak evidence in neighboring parcels. Overall, associations with affective burden were more spatially extensive and showed stronger evidence than those observed for physical burden.

**Fig. 8:**
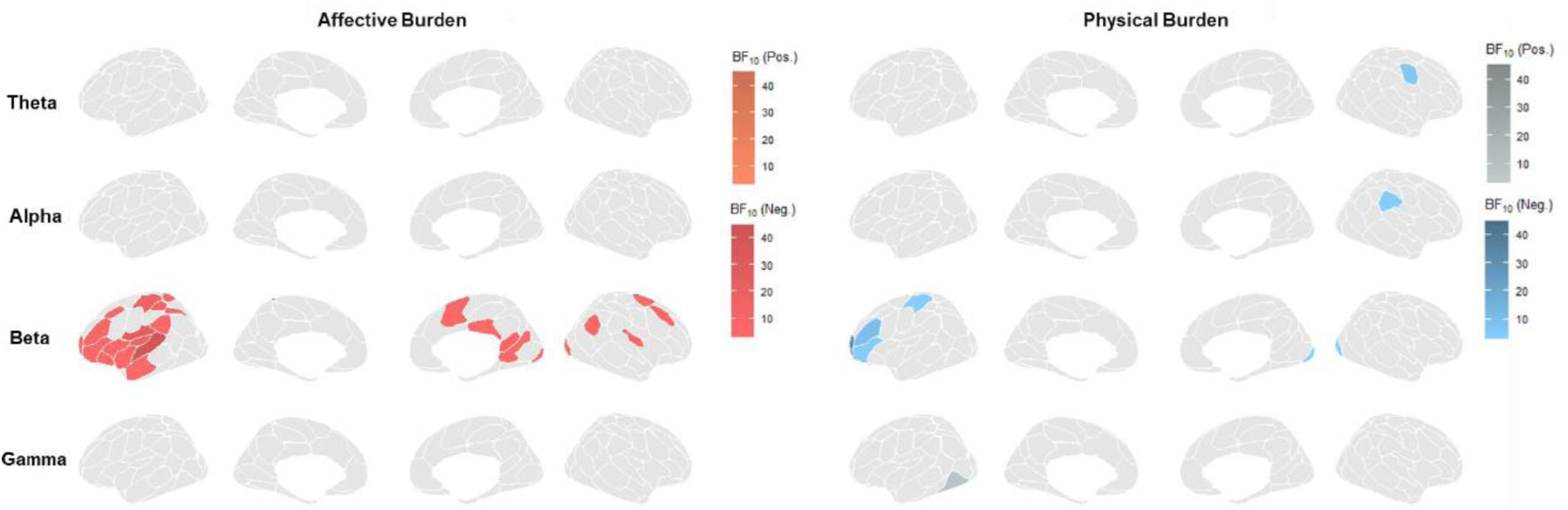
Overview of regression analyses of affective/physical burden on regional connectivity metrics, split by frequency bands. Parcellations of the Schaefer 100 parcel atlas were plotted with BFs and direction of effects. Only BFs indicating at least moderate evidence for effects are plotted. Note that for affective burden, no positive associations were plotted as a result. Connectivity was calculated as node strength of single parcels. BF = Bayes Factor, Pos. = Positive Relation, Neg. = Negative Relation.

For regional EEG power measures, including PAF, and regional connectivity in other frequency bands, we observed weak to moderate evidence against or only weak, scattered, isolated, and spatially incoherent evidence in favor of associations between EEG features and affective and physical burden

## 4. Discussion

### 4.1. Summary of Findings

In this study, we employed a data-driven, bottom-up approach to identify symptom dimensions of CP and relate them to brain function measured by EEG. This approach was motivated by the view that brain representations of CP are likely dimensional, reflecting broad symptom domains rather than singular symptoms. We identified two symptom dimensions: affective burden, which was higher in women and negatively associated with age, and physical burden, which did not differ by gender and was positively associated with age. The most robust neurophysiological finding was a negative association between affective burden and beta-band connectivity, particularly of left frontal and somatomotor regions.

### 4.2. Interpretation of Findings

Several previous studies have mapped latent psychological, social, and functional symptom dimensions of CP [7; 8; 24; 26; 39] and defined symptom-based patient subgroups [1; 12; 19; 26; 30; 35]. Results of those studies provide support for the presence of both physical and affective symptom dimensions in CP. However, findings have not been entirely consistent across studies, likely due to differences in patient-reported outcome (PRO) tools, assessed domains, and study populations. Importantly, independent psychometric analyses of the PROMIS-29 across large and clinically diverse population-based samples have consistently identified two broad latent factors reflecting physical and mental or affective health [14; 17]. The close correspondence between these established PROMIS-29 summary dimensions and the symptom dimensions identified in the present CP sample supports the construct validity of our data-driven approach. The current work extends prior PROMIS-based work by linking these dimensions to brain function and to CP. A similar approach has previously been used to associate psychopathological and personality symptom dimensions with brain connectivity patterns using fMRI in people with chronic back pain [39], underscoring the potential of linking latent symptom dimensions to brain function as a strategy for understanding the brain correlates of CP.

Regarding brain representations of identified symptom dimensions, we observed a negative association between beta-band connectivity and affective burden, primarily driven by effects in left frontal and somatomotor regions. When mapped onto the intrinsic brain networks underlying the 100-parcel Schaefer atlas [32], the strongest effects were located within the somatomotor network, with additional, less consistent effects across default mode, dorsal attention, and salience/ventral attention networks. The prominence of the somatomotor network may suggest a link between affective burden and sensorimotor integration, although this interpretation remains tentative given the distributed pattern and cross-sectional design. More broadly, the involvement of multiple large-scale networks is consistent with a network-based organization of brain function, in which broad symptom dimensions are represented across distributed systems rather than localized regions [25; 42]. Together, these findings support a dimensional view of CP in which affective symptom burden maps onto large-scale brain connectivity patterns. Notably, a recent systematic review found no consistent links between the single measure of pain intensity and EEG metrics, particularly beta connectivity. [44]. This may reflect the complex nature of CP and the limitations of simple, single-factor measures, suggesting that approaches based on empirically identified symptom dimensions could provide a more sensitive method for identifying neural links to disease severity.

### 4.3. Strengths and Limitations

Our study has several strengths. First, we applied a fully data-driven, bottom-up approach to identify symptom dimensions of CP. Unlike theory-driven frameworks, this method allows the natural structure of PROs to emerge, reducing bias in symptom selection. Second, the PROMIS-29 is brief, easy to administer, scalable, and internationally validated, facilitating replication and potential clinical implementation. Third, our EEG approach leveraged a dry-electrode system with a simple 29-channel montage and a streamlined, openly available preprocessing and analysis pipeline. This setup enables rapid, non-invasive acquisition of brain activity data, increasing feasibility for larger cohorts and potentially real-world clinical settings. Ultimately, combining behavioral and neural data provides a translational framework that links symptom dimensions to underlying brain function, offering mechanistic insights with potential clinical relevance.

Limitations apply to our study. First, caution is warranted in interpreting the connectivity results. The use of a dry 29-channel system provides relatively coarse spatial resolution, and replication with higher-density or wet-electrode systems is necessary to confirm the observed patterns. Second, the specificity of our findings remains uncertain. The symptom dimensions derived via PCA reflect the sample and questionnaire used and may not generalize to other CP populations or PRO instruments. Similarly, the EEG associations, while informative, cannot establish causal neural mechanisms. The modest sample size for the EEG subset limits the generalizability and robustness of findings, highlighting the clear need for replication in larger, independent datasets.

### 4.4. Perspectives

The integration of data-driven symptom dimensions with EEG activity illustrates a concrete pathway toward linking patient-reported outcomes to underlying neural mechanisms. While clearly needing replication, we consider our study as an example of how empirically derived symptom domains can inform mechanistic investigations, potentially guiding clinical translation. For instance, brain function patterns associated with specific symptom dimensions could be leveraged to tailor NIBS interventions, such as transcranial magnetic stimulation or neurofeedback, targeting symptom-specific circuits. Beyond NIBS, symptom-based stratification may enable prediction of individual disease trajectories, allowing clinicians to identify patients at risk for chronicity, high disability, or poor treatment response. Integrating these dimensions with longitudinal data could inform personalized treatment planning, early intervention strategies, and adaptive monitoring of therapy effectiveness.

### 4.5. Conclusion

In conclusion, using a fully data-driven approach with scalable PRO and EEG tools, we identified two symptom dimensions of CP, affective and physical burden, of which affective burden was negatively associated with beta connectivity. These results reinforce the idea that brain representations of CP reflect underlying symptom dimensions. Moreover, they demonstrate how linking latent symptom structures with neural measures can serve as a practical and scalable step toward developing clinically useful brain biomarkers for CP.

## Data Availability

The study protocol and analysis plan were preregistered (https://osf.io/7bwnh/registrations), and data and code for analyses will be made publicly available upon publication of this manuscript (https://osf.io/7bwnh/files).

## Acknowledgements

We thank Bernhard Haller for helpful comments on the statistical analysis. The authors used Grammarly and ChatGPT to improve language and readability while preparing this manuscript. The authors edited the content as needed and took full responsibility for the content of the work. The study was supported by the Deutsche Forschungsgemeinschaft (PL321/14-1, PL 321/16-1, SFB1158) and the Technical University of Munich (TUM Innovation Network *Neurotech*).

